# Oral polychemotherapy to induce remission in newly diagnosed type 2 diabetes: a pragmatic, multicenter, randomized controlled trial

**DOI:** 10.1101/2025.10.07.25337482

**Authors:** Alessandra Dei Cas, Raffaella Aldigeri, Giuseppe Maglietta, Emanuela Balestreri, Monia Garofolo, Giuseppe Daniele, Giuseppe Penno, Maddalena Trombetta, Alessandro Csermely, Monica Antonini, Angela Vazzana, Valentina Moretti, Sara Flamigni, Valentina Spigoni, Gloria Cinquegrani, Federica Fantuzzi, Marcello Monesi, Paolo Di Bartolo, Uberto Pagotto, Caterina Caminiti, Riccardo C. Bonadonna

## Abstract

**Objective:** to assess the incremental efficacy of a four-drug (POLYCHEM: metformin/pioglitazone/sitagliptin/empagliflozin) regimen vs standard diabetes care (SDC) in inducing remission in patients with newly diagnosed type 2 diabetes.

**Research Design and Methods:** A multicenter, open-label, pragmatic, phase III, randomized clinical trial was conducted in 6 Italian Diabetes Outpatient Clinics. Major inclusion criteria were age 35-75 years, HbA1c <10% (97 mmol/mol), fasting C-peptide >0.3 nmol/l, GAD-antibody negative. Patients were randomized (visit 1) to receive POLYCHEM or SDC for 16 weeks, after which, if there was regression of diabetic hyperglycemia (visit 2), drug therapy was suspended. The primary endpoint was remission (HbA1c < 6.5 %; 48 mmol/mol) assessed at least 12 weeks (visit 3) after ceasing any glucose-lowering pharmacotherapy.

**Results:** 108 patients (80% males, age: 58±9 yrs), were enrolled in the study, of whom 60 were randomized to POLYCHEM and 48 to SDC treatment. After 16 weeks, 54 (90.0%) and 37 (77.1%) patients achieved regression of diabetic hyperglycemia in the POLYCHEM and SDC arm, respectively (p=0.002). At visit 3, 23 (38.3%) and 21 (43.8%) patients achieved diabetes remission in the POLYCHEM and SDC arm, respectively (p=0.59). The difference between the upper 95% CI in POLYCHEM (50.9%) and the lower 95% CI in SDC (30.4%) of diabetes remission was 20.5%.

**Conclusion:** A four-drug regimen with minimal to null weight-lowering effect is not superior to current SDC in achieving remission in patients with newly diagnosed type 2 diabetes. If a superiority of POLYCHEM over SDC exists, it is expected to be less than 21%.

**Trial registration:** ClinicalTrials.gov number: NCT04271189

## 1. Introduction

Diabetes affects over 600 million adults worldwide, and projections indicate a further rise to 853 million (13%) by 2050 [1]. his epidemic, largely driven by westernized lifestyles and obesity, is associated with severe complications and reduced quality of life, and contributes to nearly one in ten deaths worldwide. Beyond clinical care diabetes already accounts for 12% of global health expenditure, placing a heavy burden on healthcare systems and societies. Within this scenario type 2 diabetes (T2D) constitutes over 90% of all cases, underscoring the urgency of developing innovative, scalable and sustainable strategies for prevention, management and - critically – remission [2]. Remission, as defined by the latest consensus criteria as HbA1c to < 6.5% over a period of at least 3 months in absence of glucose-lowering pharmacotherapy or at least 6 months after the lifestyle intervention [3,4], has emerged as a clinically relevant and doable therapeutic goal. When achieved early, remission can delay or even prevent T2D-related complications, with benefits comparable to preventive strategies, while substantially reducing disease burden. To date, metabolic surgery [5,6] and (very) low-calorie diets (VLCDs) [7,8,9] have consistently been shown to induce sustained T2D remission. These approaches have demonstrated that remission is achievable through substantial body weight reductions (>15–20%) [10 11,12]. More recently, comparable weight loss obtained through treatment with Glucagon-like peptide 1 (GLP-1) receptor agonists [13], as well as dual (GLP-1 receptor and Glucose-dependent Insulinotropic Polypeptide-GIP-receptors) [14] and triple (GLP-1, GIP and glucagon receptors) [15] agonists, has been shown to reduce HbA1c levels ≤6.5% (48 mmol/mol) in a high proportion of patients (50-80%). However, upon treatment discontinuation, body weight tends to rebound and glycemic control deteriorates, indicating that remission is pharmacologically sustained rather than permanent.

Other interventions aiming at diabetes remission have also been linked to significant weight loss. An intensive strategy combining lifestyle intervention, metformin and insulin glargine/lixisenatide achieved higher remission rates than standard care [16]. In contrast, a 16-wk intervention with insulin degludec/liraglutide failed to reach a statistically significant improvement in rates of diabetes remission vs standard care, but this difference paralleled a minimal weight loss difference (<2 kg) [17].

As to weight-loss independent interventions, in newly-diagnosed individuals with T2D, the initiation of insulin therapy reportedly induced remission, apparently by restoring beta-cell secretory function, presumably through glucose normalization and reduction in glucotoxicity [18]. Interventions with oral anti-diabetes drugs featuring glucose, but modest or no weight-lowering, action have provided mixed results [19,20]. However, none of these approaches has tested the simultaneous use of four agents targeting distinct aspects of T2D pathophysiology at its early stage. In this context, we conducted a proof-of-concept study to assess whether a four-drug regimen, engaging multiple glucose-lowering mechanisms, could induce regression of diabetic hyperglycemia (HbA1c <6.5%, 48 mmol/mol) and, after 3-month treatment suspension, achieve diabetes remission in a clinically meaningful higher proportion of patients compared with standard diabetes care (SDC) in newly diagnosed T2D.

## 2. Methods

### 2.1 Study design

This is a multicenter, open label, pragmatic, phase III, randomized 1:1 controlled clinical trial, designed to compare the efficacy of early initiation of a new therapeutic strategy of four drugs (POLYCHEM) vs conventional care (SDC), in subjects newly diagnosed type 2 diabetes (NCT04271189). The study involved six Italian Diabetes Outpatient Clinics (Parma, Ferrara, Ravenna, Bologna, Verona and Pisa). The study was conducted according to the guidelines of the Declaration of Helsinki and approved by the local Ethics Committee “Comitato Etico dell’Area Vasta Emilia Nord” with EUDRACT registration no. 2018-000833-12. Written informed consent was obtained from all subjects involved in the study. The planned duration of the study was two years, but an extension was granted due to the COVID-19 pandemic. Therefore, the enrolment period spanned from October 2020 to July 2023. The protocol was amended in January 2022, after the publication of the new consensus report which revised remission criteria [4]. These new criteria significantly differed from the previous ones, and included the achievement of HbA1c <6.5% (<48 mmol/mol) for 12 weeks, in the absence of any glucose lowering pharmacologic treatment [4]. In response to the updated definition and to operational requirements, the study design was modified from the initially planned two-phase adaptive approach (phase IIb-phase III) with 2:1 ratio (POLYCHEM:SDC) to a phase III design with 1:1 allocation ratio (amendment approved on 7 April 2022).

### 2.2 Study population

Inclusion criteria included age ≥ 35 and ≤ 75 years, newly diagnosed (within 6 months) type 2 diabetes according to American Diabetes Association (ADA) criteria [21], negative anti-GAD antibodies, C-peptide ≥0.3 nmol/l, body mass index (BMI) ≥23 and ≤40 Kg/m^2^, HbA1c ≤10% (86 mmol/mol), absence of diabetic retinopathy. Main exclusion criteria were: current or history of cancer within the last 5 years, acute cardiovascular event in the previous six months, Class III-IV heart failure according to the New York Heart Association (NYHA) classification, corticosteroid-based drugs or immunosuppressive medications, estimated creatinine-clearance (eGFR) <45 mL/min/1.73 m^2^ assessed by with CKD-EPI formula, severe organ failure, multi-injection insulin therapy, contraindications to any of the study drug, as well as pregnancy or lactation. Women of childbearing age were eligible only if on birth control. The enrolment period spanned from October 2020 to July 2023.

### 2.3 Study intervention

Patients were randomized 1:1 to either SDC or “POLYCHEM” (intervention arm). The latter consisted in the combination of four drugs: metformin, extended-release formulation, titrated to 2000 mg/day, pioglitazone 15 mg/day, sitagliptin 100 mg/day, and empagliflozin 10 mg/day. SDC consisted in the conventional treatments provided at each center [20].

Patients with newly diagnosed type 2 diabetes were randomized at baseline (V1) to receive POLYCHEM or SDC for 16 weeks. At the end of this period (V2), based on the original protocol version (simultaneous nondiabetic values of fasting glucose<126 mg/ml, 2-hour glucose after OGTT <200 mg/ml and HbA1c <6.5%) or according to the amended protocol [HbA1c < 6.5% (<48 mmol/mol)]) diabetic hyperglycemia was assessed. If regression of diabetic hyperglycemia [HbA1c < 6.5% (<48 mmol/mol)] was achieved, anti-diabetes therapy was discontinued irrespective of the randomization arm and patients were monitored for an additional 12 weeks on no pharmacological therapy (V3). Conversely, if diabetic hyperglycemia regression was not achieved at V2, anti-diabetes therapy was modified irrespective of the randomization arm, according to clinical guidelines at the discretion of the treating physician. Furthermore, at V2, investigators could decide to stop the POLYCHEM treatment at any time at their discretion, for safety or intolerability reasons, based on laboratory parameters or clinical events. At 28 weeks (V3) HbA1c was re-assessed, to test diabetes remission [<6.5% (48 mmol/mol)] after ceasing any glucose-lowering pharmacotherapy for 12 weeks.

### 2.4 Objectives and endpoints

The primary objective was to determine whether POLYCHEM was superior to conventional care in inducing diabetes remission defined as HbA1c <6.5% (48 mmol/mol) for at least three months after cessation of any glucose-lowering intervention (2021 Consensus Report) [3] which substantially revised the 2009 criteria [4]. The primary endpoint is the difference in diabetes remission rates (HbA1c < 6.5% (48 mmol/mol) assessed at least 12 weeks (V3) after ceasing any glucose-lowering pharmacotherapy between study groups. Secondary endpoints include hyperglycemic regression (at 16 weeks) and changes in HbA1c and body weight throughout the study.

Planned secondary objectives, including quality of life and direct costs [see clinicaltrials.gov registration NCT04271189], are not addressed in this paper and will be reported in a further publication.

### 2.5 Sample size

To evaluate the efficacy of POLYCHEM in terms of percentage of remissions at seven months, sample size was determined by using 5% one-sided lower limit confidence intervals for the difference between two proportion test. The difference between the two groups was defined at 15% and the lower limit of its confidence interval must not be smaller than 5%. Therefore, 92 patients for each group (total n=184) were necessary. However, owing to the end of funding, only 108 patients were randomized and, since the study initially was conducted according to a two-phase adaptive approach (phase IIb-phase III) with 2:1 ratio (POLYCHEM:SDC) and only subsequently amended into a phase III design with 1:1 allocation ratio (see Study Design), the 1:1 randomization ratio was not achieved (see Results). The post-hoc power calculation of the study, given the number of patients actually randomized and the proportion of diabetes remission achieved in the SDC arm (43.8%), yielded a minimum detectable difference of 25% at p < 0.05 in the proportion of diabetes remission between POLYCHEM and SDC arms with power = 0.80.

The primary efficacy analysis was conducted based on Intention-To-Treat (ITT) principles. Univariable and multivariable logistic regression were implemented to describe the association between the primary endpoint and treatment, as well as other predictors (taken from the literature or with p value <0.1 in the univariable analysis).

### 2.6 Randomization and Blinding

Random allocation sequences were generated stratifying per center and using blocks of 2 or 3. Randomization lists were generated by the trial statistician by specifying a unique seed for each list and using the blockrand R package. Allocation concealment was ensured by storing lists in the centralized electronic Case Report Form (eCRF) developed by an external service using the OpenClinica platform. Blinding was not possible due to the process of treatment administration and clinical outcome assessment.

### 2.7 Statistical analysis

The analyses were conducted according to ITT. Primary efficacy endpoint was analyzed using a two-proportion chi-square test to compare remission proportions in the two arms. Baseline characteristics are presented as median (Interquartile Range, IQR) and mean (Standard Deviation, SD) for quantitative variables and absolute and relative frequencies for qualitative variables. Missing data were not imputed. Mixed model for repeated measures were performed to estimate variable changes in time between the two treatment arms. Univariate and multivariate logistic regression analysis were performed for remission rate. All statistical analyses were performed with R Statistical Software, V 4.3.0 and SPSS (IBM Statistics v.29).

## 3. Results

### 3.1 Study population

Overall, between October 2020 and July 2023, 110 patients with newly-diagnosed type 2 diabetes were screened, of whom 108 (98.2%) were randomized, 60 (55.5%) to the POLYCHEM intervention and 48 (44.5%) to the SDC control arm (Supplementary Figure 1). Since the patient randomization was initially 2:1, a higher proportion of patients was randomized to the intervention than to the standard care arm.

Two patients in the SDC arm (4.2%), and 2 (3.3%) in the POLYCHEM arm were lost at follow-up. Table 1 shows baseline demographic and clinical characteristics of participants which were similar between the two groups, as expected by the randomization procedure. Overall, patients were prevalently male (80%), with median age 58 years. Mean BMI was 29.9 Kg/m^2^ and baseline HbA1c was 6.6% (49mmol/mol); almost 18% were active smokers.

**Table 1.** Baseline Characteristics of the study population.

|  | **Overall, N=108** | **POLYCHEM, N = 60** | **SDC, N = 48** |
| --- | --- | --- | --- |
| **Age (years)** | 58 ± 9 | 60 ± 8 | 56 ± 9 |
| **Female sex** | 22 (20%) | 13 (22%) | 9 (19%) |
| **Ethnicity, caucasian** | 108(100%) | 60(100%) | 48(100%) |
| **BMI (kg/m<sup>2</sup>)** | 29.9± 4.8 | 29.6±5.3 | 30.3± 4.1 |
| **SBP (mmHg)** | 134±14 | 135±14 | 133±15 |
| **DBP (mmHg)** | 82±11 | 82±11 | 83±11 |
| **Waist circumference(cm)** | 105.0±11.5 | 104.9±11.0 | 104.9±12.2 |
| **HbA1c (mmol/mol)** | 49(44-55) | 48(44-56) | 49(43-55) |
| **HbA1c (%)** | 6.6(6.2-7.2) | 6.5(6.2-7.3) | 6.6(6.1-7.2) |
| **Fasting plasma glucose (mg/ml)** | 125(114-138) | 124(110-140) | 126(116-137) |
| **Fasting plasma glucose (mmol/L)** | 6.94(6.33-7.67) | 6.89(6.11-7.78) | 7.00(6.44-7.61) |
| **Total cholesterol (mg/ml)** | 175.6±38.7 | 179.4±36.2 | 170.8±41.6 |
| **HDL (mg/ml)** | 47.6±10.9 | 47.8±11.2 | 47.5±10.6 |
| **LDL (mg/ml)** | 101.7±34.1 | 103.7±31.8 | 99.1±37 |
| **TG (mg/ml)** | 119.5(85.5-155.5) | 131.5(89-157) | 110(79.7-147.5) |
| **AST (U/L)** | 23(20-28.5) | 23(19-28) | 24(20-32) |
| **ALT (U/L)** | 29(21-40) | 29(22.2-38) | 26(20-41) |
| **Lipase (U/L)** | 34(22-45) | 36(26-48.5) | 31(20-44) |
| **eGFR (ml/min)** | 92.9±15.3 | 92.1±16.9 | 93.9±13.1 |
| **ACR (mg/gr)** | 9.1(4.2-20) | 10(4.4-38.9) | 7.3(4.1-13.3) |
| **Current Smoker** | 19 (17.6%) | 8 (13.3%) | 11 (22.9%) |

Data are given as n (%) for categorical variables and as mean ± SD or median (interquartile range) for continuous variables. Abbreviations HbA1c= glycated hemoglobin A1c, BMI= Body-Mass Index, SBP= Systolic blood pressure, DBP= Diastolic blood pressure, HDL-C= high-density lipoprotein-cholesterol, LDL-C= low-density lipoprotein-cholesterol, TG= triglycerides, AST= Aspartate Aminotransferase, ALT= Alanine Aminotransferase, eGFR= eGFR, estimated glomerular filtration rate, ACR= albumin to creatinine ratio.

### 3.2 Anti-diabetes therapy in the SDC arm

Glucose lowering drugs in the SDC arm at baseline (V1) and during follow-up (V2-V3) are reported in Supplementary Table 1. A total of 5 patients (12.5%) were treated with basal insulin therapy and 4 (10%) with GLP-1 receptor agonist.

### 3.3 Effect of the POLYCHEM on diabetes remission (primary endpoint)

The POLYCHEM intervention exerted no significant effects on the rate of diabetes remission in newly diagnosed patients at weeks 28 as shown in figure 1A. At weeks 28 (V3) 38.3% (95% C.I.: 26.8 – 50.9) of the individuals in the POLYCHEM arm compared to 43.8% (95% C.I.: 30.4 – 57.8) in the SDC arm achieved the primary endpoint (p=0.59). The difference between the upper 95% C.I. of POLYCHEM (50.9%) vs the lower 95% C.I. of SDC (30.4%) was 20.5%.

**Figure 1.**
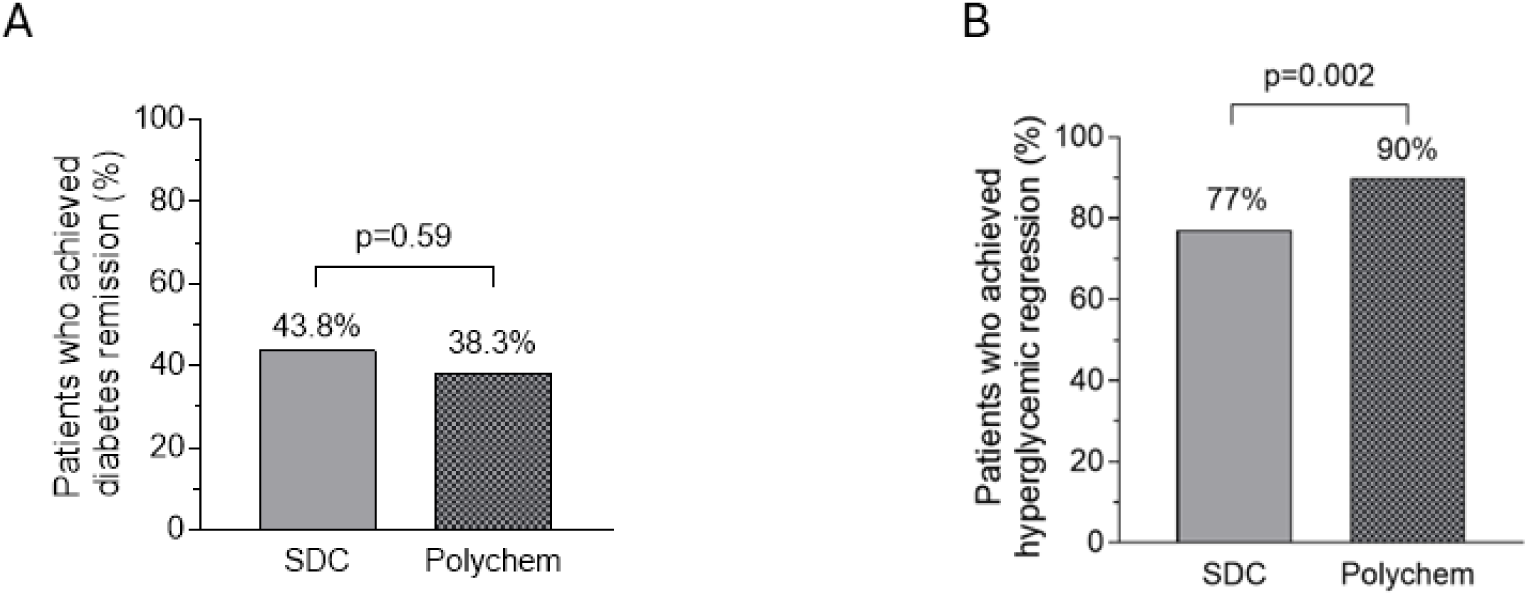
A. Patients (%) who achieved diabetes remission at 28 weeks in the intervention arm vs SDC arm. B. Patients (%) who achieved hyperglycemia regression at 28 weeks in the intervention arm vs SDC arm.

### 3.4 Effect of the POLYCHEM in the diabetic hyperglycemia regression

More patients achieved diabetic hyperglycemia regression (HbA1c < 48 mmol/mol at 16 weeks) in the POLYCHEM arm (90%) than int SDC (77%) arm (p=0.002), as shown in figure 1B. Supplementary Table 2 shows the percentages of patients who achieves diabetic hyperglycemia regression at 16 weeks and diabetes remission at 28 weeks (HbA1c <48 mmol/mol after treatment discontinuation) [4]. Of note, 19 (31.6%) individuals in the POLYCHEM and 11 (22.9%) in the SDC arm achieving hyperglycemia regression continued anti-diabetes therapy due to the remission criteria used before protocol amendment which required the fulfilment of not only HbA1c <6.5% (48 mmol/mol) levels but also the simultaneous normalization of fasting plasma glucose <126 mg/dl (7 mmol/l) and 2-hour glucose levels <140 mg/dl (7.8 mmol/l) following a glucose tolerance test 75g.

### 3.5 Effect of the POLYCHEM on HbA1C and body weight

Figure 2 depicts changes in HbA1c (A) and body weight (B) during the study period in the two arms. At randomization, mean HbA1c values were identical in the two groups (6.6%; 49.7 mmol/mol) and decreased over the first 16 weeks (V2) in both arms, with a non-significant difference in favor of the POLYCHEM arm (Δ-1.8, p =0.24). At the second follow-up visit (28 weeks) (V3), HbA1c gradually increased in both arms, significantly more in the POLYCHEM arm (Δ+4.8 p<0.001). In the GLM model a significant time decrease in HbA1c was present (p<0.001) without treatment effect (p=0.93). At baseline (V1), body weight was slightly, but not significantly higher in the SDC arm (+1.9 Kg; p=0.53), at 16 weeks body weight decreased in both arms without significant differences (p=0.21), at 28 weeks (V3) a significant increase in body weight was observed in the POLYCHEM arm vs SDC (+1.28 Kg; p=0.02). Similarly, in the GLM model a significant time*treatment effect was observed (p<0.02) with a significantly greater reduction of weight in subjects randomized to SDC as compared to POLYCHEM arm.

**Figure 2.**
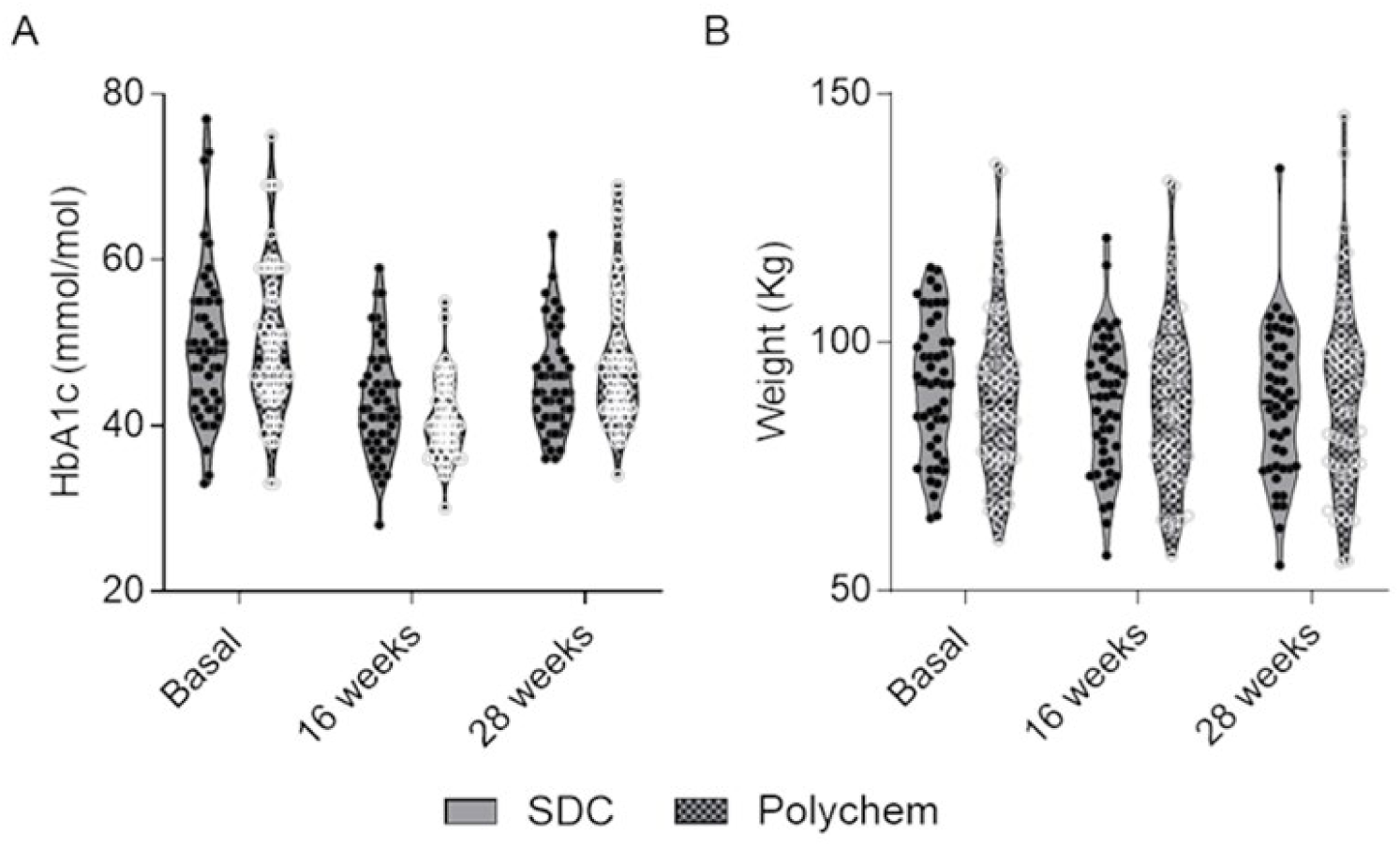
A. HbA1c (mmol/mol) and B. Body weight (kg) values in the two arms at baseline and during follow-up

Changes in glucose and other clinical parameters at different time points during the study are shown in Supplementary Table 3. No significant differences were seen between treatment arms.

### 3.6 Variables associated to diabetes remission

In the univariable logistic regression analysis, in the whole population, remission rates were negatively associated with higher baseline HbA1c (OR=0.92 [95%CI: 0.87-0.97]; p=0.002) and fasting plasma glucose (FPG) levels (OR=0.94 [95%CI: 0.91-0.97]; p<0.001) and positively associated with a higher decrease in body weight at 16 weeks (OR=1.14 [95%CI: 1.02-1.28]; p=0.02). In a multivariate analysis, after adjusting for age, gender, baseline HbA1c values and weight reduction after 16 weeks, only lower HbA1c values (OR= 0.91 [95%CI 0.86-0.97]; p=0.002) and weight loss (OR 1.15 (95%CI 1.03-1.29]; p=0.01) remained independent predictors of diabetes remission at 28 weeks in the whole population. When the analysis was conducted separately per randomization arm, in the POLYCHEM arm, baseline HbA1c values (OR=0.83 [95%CI 0.73-0.94] p=0.002) and weight reduction after 16 weeks (OR=1.29 [95%CI 1.04-1.60] p=0.002) were confirmed independent predictors of DM remission, whereas in the SDC arm only age [OR=0.92 (95%CI 0.85-0.99) p=0.04] was independently associated with the primary endpoint (data not shown).

### 3.7 Adherence to therapy

The mean percentage of compliance to the POLYCHEM therapy was 96.1% (SD=7.9%). Two subjects (3.3%) randomized to POLYCHEM did not take therapy because of intolerance (non-specific symptoms such as such as fatigue and malaise). In the SDC arm (Table 2) three subjects (6.3%) decreased the dosage of metformin due to gastrointestinal side effects.

**Table 2.** Distribution of adverse events (AE) in study participants.

| **Overall (n, %)** | **POLYCHEM (n, %)** | **SDC (n, %)** | **p-value** |
| --- | --- | --- | --- |
| **40, 37%** | 25,41.7% | 15,31.3% | 0.26 |
| **Severity** |  |  |  |
| -mild | 21, 84% | 11, 73% | 0.24 |
| -moderate | 4, 16% | 1, 7% |  |
| -severe | 0 | 3, 20% |  |
| **Causality** |  |  |  |
| -not correlated | 16, 64% | 12, 80% | 0.33 |
| -possible/probable | 9, 36% | 3, 20% |  |
| **Type of AE:** |  |  |  |
| -Gastrointestinal | 12, 20% | 5, 10% | 0.36 |
| -Urogenital | 7, 12% | 4, 8% | 0.93 |
| -SARS-COV2 infection | 3, 5% | 3, 6% | 0.49 |
| -Respiratory tract infection | 6, 10% | 0 | 0.04 |
| -Dizziness | 1, 2% | 3, 6% | 0.10 |
| -Low back pain | 2, 3% | 2, 4% | 0.60 |
| -Paresthesia | 1, 2% | 1, 2% | 0.70 |
| -Hypertension | 1, 2% | 1, 2% | 0.70 |

Data are given as n, (%).

### 3.8 Adverse events

Over the 28-weeks’ study period, 40 patients, 25 in POLYCHEM (41.7%) and 15 in SDC (31.3%) experienced adverse events, as summarised in Table 2, without significant differences. Three serious adverse events (SAEs) occurred in the SDC arm and included prostate cancer, acute cystopyelitis and hospitalization for road accident. The most common reported events were gastrointestinal (n=17), urogenital (n=11), SARS-COV2 infections (n=6), and respiratory infection with or without fever (n=6). Most of them were classified as mild AEs (n=32/40, 80%) and not related to experimental drugs (n=28/40, 70%). In the POLYCHEM arm 6 subjects (10%) reduced the dosage of metformin due to gastrointestinal side effects, 3 (5%) discontinued empagliflozin because of urogenital infections while 2 (3.3%) permanently discontinued all drugs.

## 4. Discussion

Given the growing interest in diabetes remission as a therapeutic goal, this proof-of-concept trial was designed to test whether a fully oral polypharmacological regimen (sitagliptin + metformin + empagliflozin + pioglitazone) administered over 16 weeks was superior to standard diabetes care (SDC) in achieving remission in newly diagnosed type 2 diabetes. Remission rates were comparable between the two groups (38.3% in POLYCHEM vs 43.8% in SDC), and no statistically significant differences in body weight were observed. These results suggest that pharmacologically induced remission is feasible through weight-independent mechanisms, although the POLYCHEM strategy did not demonstrate superiority over SDC.

We had designed the study with the goal of being able to capture an absolute difference in the proportion of diabetic remission of 15%. Owing to end of funding, the actual power of this study was an 80% chance of detecting an absolute difference of 25% in the proportion of diabetes remission at p < 0.05. Since the proportion of diabetes remission in the SDC arm was 43.8%, POLYCHEM should have induced almost twice more (68.8%) of diabetes remissions than it actually did (38.3%) for reaching statistical significance. Furthermore, the upper 95% confidence interval of the proportion of diabetes remission in the POLYCHEM arm was 50.9%, much lower than the value (68.8%) which would be expected to be statistically significant. Finally, and most importantly, the difference between the upper 95% C.I. of POLYCHEM and the lower 95% C.I. of SDC in diabetes remission was 20.5%. Thus, we think that this study provides evidence that the superiority, if any, of POLYCHEM over SDC in inducing diabetes remission is less than 21%.

Dietary interventions [7,8,9] and bariatric surgery [22,23,24,25] have consistently proven effective in inducing durable diabetes remission, with weight loss serving as the common euglycemic mechanism across these different strategies. In bariatric surgery, additional biological factors - including changes in gut microbiota, bile acid metabolism, and the secretion of appetite-modulating peptide hormones-may further contribute to remission [26]. In contrast, remission in our trial occurred without clinically meaningful (i.e. ≥ 5%) weight reduction, suggesting the existence of alternative mechanisms of metabolic recovery.

Beyond lifestyle and surgical interventions, early intensive pharmacological strategies at diagnosis have also been investigated. Early intensive insulin therapy in patients with recent-onset type 2 diabetes showed durable improvements in β-cell function [27,28,29] and led to prolonged glycemic remission compared with treatment with oral anti-diabetes drugs. Insulin therapy, by rapidly correcting hyperglycaemia and glucotoxicity, can restore beta-cell secretory activity. More recently, the combination of insulin pump therapy with Low-Calorie Diet (LCD) intervention was associated with improved remission rates in patients with newly diagnosed type 2 diabetes [30].

The Remission Evaluation of Metabolic Intervention in Type 2 Diabetes (REMIT) pilot trial showed that a polypharmacological approach over 16 weeks (metformin + acarbose + insulin glargine) was more effective in inducing normoglycemia and remission compared to standard care (41% vs. 14%) [31]. Similarly, the REMIT-dapa trial [32] and REMIT-sita trial [33] evaluated the efficacy of polypharmacological regimens with insulin glargine + metformin + dapagliflozin and insulin glargine + metformin/sitagliptin, respectively, *vs* standard care. Both regimens, maintained for 12 weeks, and both demonstrated superior remission rates [HbA1c ≤6.5% (48 mmol/mol)] and reduced relapse compared to standard care.

To our knowledge, our study is the first randomized trial testing a fully oral, multi-target regimen in newly diagnosed type 2 diabetes using formal remission criteria. Compared with previous studies on early intensive insulin therapy and polypharmacological regimens that included insulin, our proof-of-concept uniquely addressed whether remission could be achieved through fully oral strategies not associated with significant weight loss.

While remission rates in our study were similar between the intervention arm compared to the SDC arms (38.3% *vs* 43.8%), the overall remission frequency in both arms (≈40%) was markedly higher than the spontaneous remission typically reported in newly diagnosed type 2 diabetes (≈4–6%) ^[^34], suggesting that early pharmacological intensification provided in outpatient specialist clinics may be a viable strategy to reverse hyperglycemia.

The potential mechanisms underlying remission in our study deserve specific consideration. Unlike weight-dependent interventions, the POLYCHEM regimen simultaneously targeted the key underlying mechanisms driving the onset and progression of type 2 diabetes, namely insulin resistance, β-cell dysfunction, and glucotoxicity. Of note, the current standard therapeutic paradigm has shifted from stepwise intensification to earlier initiation of combination therapies, including injectables, at the time of diagnosis. Such a multimodal approach may enhance β-cell preservation and improve insulin sensitivity independently of body weight even in the current standard care. A possible explanation for the comparable remission rates between groups, indeed, is that in the SDC arm many patients also received early multidrug therapy in line with contemporary treatment guidelines, thereby narrowing the difference in the treatment strategies. Additionally, the therapeutic strategies that have so far achieved diabetes remission have largely relied on substantial weight loss, which remains one of the strongest predictors of remission, as demonstrated by interventions such as (very) low-calorie diets [35] and bariatric surgery [36]. In contrast, the rationale of our study was to investigate whether remission could be achieved through pharmacological agents that do not promote significant weight loss, but rather target the underlying pathophysiological mechanisms present at diabetes onset, with the potential to restore metabolic function and reverse hyperglycaemia.

Overall, in our study, no statistically significant differences in body weight were observed between the two treatment groups, underscoring that pharmacologically induced remission can occur via weight-independent mechanisms. At 28 weeks, body weight slightly decreased in the standard care group, whereas patients in the POLYCHEM group showed a mean increase of +1.2 kg from baseline. This differential effect may have attenuated the efficacy of the POLYCHEM regimen and was confirmed in multivariable analysis, where weight loss emerged as an independent predictor of remission. Indeed, greater weight loss was an independent “predictor” of diabetes remission in the multivariable analysis.

The POLYCHEM strategy was well tolerated, supporting its potential as a viable therapeutic approach to be further evaluated and implemented in larger population-based studies. Nevertheless, the study did not demonstrate superiority of a 16-week, four-drug oral regimen over SDC in inducing remission, and the findings should be interpreted as exploratory. Furthermore, the overall evidence of this study suggests that the putative, but quite unproven, superiority of POLYCHEM over SDC must be a figure of absolute difference of diabetes remission less than 21%.

Our study presents some strengths namely the randomized design and the rigorous assessment of diabetes remission based on the standardized criteria established by international consensus statements and clinical guidelines. This contrasts with recent incretin-based trials [13,14,15], where HbA1c ≤6.5% was reported under ongoing pharmacological treatment, thereby not fulfilling remission definitions [4].

Limitations include the relatively small sample size and the absence of a predefined therapeutic algorithm in the SDC group, which may have introduced heterogeneity and influenced outcome comparisons.

## 5. Conclusions

In conclusion, this randomized trial shows that a fully oral, insulin-sparing polypharmacological regimen can induce remission in a substantial proportion of patients with newly diagnosed T2D, independent of weight loss. However, it did not prove superior to standard care. These findings support early pharmacological intensification as a potential avenue for remission, but larger and longer-term trials are needed to define the role of oral polypharmacological regimens in the evolving therapeutic landscape of type 2 diabetes.

## Author contributions

Riccardo C. Bonadonna: Writing – original draft, Visualization, Methodology, Investigation, Funding acquisition, Formal analysis, Data curation, Conceptualization, Supervision. Alessandra Dei Cas: Writing – original draft, Visualization, Methodology, Investigation, Formal analysis, Data curation, Supervision. Raffaella Aldigeri: project administration, data curation, formal analysis, writing-review & editing. Emanuela Balestreri: investigation, writing-review & editing. Monica Antonini: investigation, writing-review & editing. Angela Vazzana: investigation, writing-review & editing. Valentina Moretti: investigation, writing-review & editing. Monia Garofolo: investigation, writing-review & editing. Giuseppe Daniele: investigation, writing-review & editing. Giuseppe Penno: investigation, writing-review & editing Maddalena Trombetta: investigation, writing-review & editing. Alessandro Csermely: investigation, writing-review & editing. Marcello Monesi: investigation, writing-review & editing. Sara Flamigni: investigation, writing-review & editing. Paolo Di Bartolo: investigation, writing-review & editing. Uberto Pagotto: investigation, writing-review & editing. Giuseppe Maglietta: data curation, formal analysis. Valentina Spigoni: investigation and resources. Gloria Cinquegrani: investigation and resources. Federica Fantuzzi: investigation and resources. Caterina Caminiti: writing-review & editing.

## Data Availability

All data produced in the present studyare not publicly available but are available from the corresponding author on reasonable request

## Data availability

Some or all datasets generated during and/or analyzed during the current study are not publicly available but are available from the corresponding author on reasonable request.

