## Supplementary material for "Oral polychemotherapy to induce remission in newly diagnosed type 2 diabetes: a pragmatic, multicenter, randomized controlled trial"

CLINICAL SITES

•	Endocrinology and Metabolic Diseases, Department of Medicine and Surgery, University of Parma, Parma, Italy, site PI and Co-PI Prof. Riccardo Bonadonna, Prof. Alessandra Dei Cas

•	Division of Endocrinology, Diabetes and Metabolism, Department of Medicine, University of Verona, Verona, Italy, site PI Prof. Maddalena Trombetta

•	Division of Metabolic Diseases and Diabetes, Department of Clinical and Experimental Medicine, University of Pisa, Pisa, Italy: site PI Prof. Giuseppe Penno

•	Local Health Authority of Romagna, Ravenna, Italy: site PI Dr. Paolo Di Bartolo

•	Division of Endocrinology and Diabetes Prevention and Care, IRCCS Azienda Ospedaliero-Universitaria di Bologna, Department of Medical and Surgical Sciences, Alma Mater Studiorum University of Bologna, Bologna, Italy, site PI Prof. Uberto Pagotto

•	Diabetes Unit, Primary Care Department, AUSL Ferrara: site PI Dr. Marcello Monesi

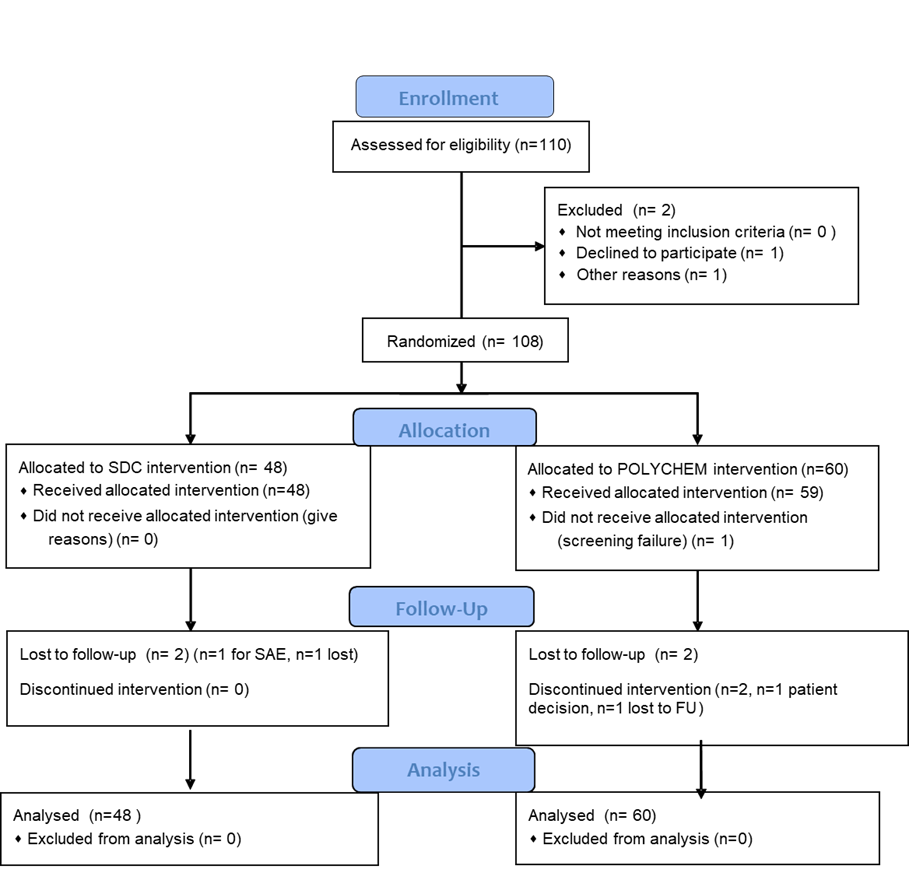

Supplemental Figure 1. Study flow diagram according to ITT analysis

| **N of subjects(%)** | **Baseline (V1)** | **16 weeks (V2)** | **28 weeks (V3)** |
| --- | --- | --- | --- |
| **Glucose lowering drugs** |  |  |  |
| **None** | 2 (4.2%) | 26 (54.2%) | 26 (43.8%) |
| **1 oral** | 22 (45.8%) | 8 (16.7%) | 7 (14.6%) |
| **2 orals** | 14 (29.2%) | 8 (16.7%) | 8 (16.7%) |
| **3 orals** | 1 (2.1%) | 2 (4.2%) | 2 (4.2%) |
| **1 oral + 1 subcutaneous** | 6 (12.5%) | 3 (6.2%) | 3 (6.2%) |
| **2 orals + 1 subcutaneous** | 3 (6.2%) | 1 (2.1%) | 1 (2.1%) |
| **1 subcutaneous** | 0 | 0 | 1(2.1%) |

Supplemental Table 1. Glucose lowering drug in the SDC arm at baseline and during follow up. Data are expressed as n (%)

| **Arm** | **Diabetic Hyperlycaemia regression: patients with HbA1c <48 mmol/mol at 16 weeks** | **Patients who discontinued antidiabetic treatments during week 16-28** | **Patients who maintained HbA1c <48 mmol/mol independently of anti-diabetes therapy cessation** | **DIABETES REMISSION at 28 weeks** |
| --- | --- | --- | --- | --- |
| **POLYCHEM** | 54/60 | 35/60 | 35/60 | 23/60 |
|  | 90.00% | 58.37% | 58.33% | 38.33% |
| **SDC** | 37/48 | 26/48 | 33/48 | 21/48 |
|  | 77.08% | 54.2% | 68.75% | 43.75% |

Supplemental Table 2. Percentages of patients who achieved the primary endpoint and the three relative necessary conditions.

|  | POLYCHEM | | | SDC | | | p-val |
| --- | --- | --- | --- | --- | --- | --- | --- |
|  | 0 | 16 w | 28 w | 0 | 16 w | 28 w |  |
|  | n=60 | n=57 | n=57 | n=48 | n=48 | n=46 |  |
| HbA1c (mmol/mol) | 48(44-56) | 40(38-44) | 46(42-52) | 49.5(42.7-55.0) | 42(38-47.2) | 44(41-48.2) | 0.94 |
| FPG (mg/dL) | 124(110-144) | 113(104.4-129) | 127(108-140) | 126(116-137.2) | 118(100.7-130.2) | 122(108.7-1327) | 0.64 |
| Weight (kg) | 89.1±16.6 | 87.7±16.7 | 89.6±18.1 | 91.0±13.9 | 87.6±13.6 | 87.6±14.9 | 0.81 |
| BMI (kg/m^2^) | 29.6±5.23 | 29.5±3.75 | 30.0±4.28 | 30.3±4.11 | 29.2±3.93 | 29.2±4.36 | 0.77 |
| Waist  circumference (cm) | 105±11.0 | 103±10.8 | 104±11.7 | 105±12.2 | 101±17.9 | 102±12.3 | 0.46 |
| SBP (mmHg) | 135±14.1 | 131±15.9 | 135±15.7 | 133±14.8 | 132±13.4 | 131±14.0 | 0.43 |
| DBP (mmHg) | 82.2±10.7 | 82.0±12.1 | 82.8±10.3 | 82.8±10.6 | 83.5±8.9 | 83.6±8.7 | 0.65 |

Supplemental Table 3. Glucose and clinical parameters at different time visits. Data are expressed as mean ± SD if normally distributed, or median (IQR) if skewed. P-value from generalized linear model for repeated measures.
